# Delayed cortical engagement associated with balance dysfunction after stroke

**DOI:** 10.1101/2023.11.28.23299035

**Authors:** Jacqueline A. Palmer, Aiden M. Payne, Jasmine L. Mirdamadi, Lena H. Ting, Michael R. Borich

## Abstract

Cortical resources are typically engaged for balance and mobility in older adults, but these resources are impaired post-stroke. Although slowed balance and mobility after stroke have been well-characterized, the effects of unilateral cortical lesions due to stroke on neuromechanical control of balance is poorly understood. Our central hypothesis is that stroke impairs the ability to rapidly and effectively engage the cerebral cortex during balance and mobility behaviors, resulting in asymmetrical contributions of each limb to balance control. Using electroencephalography (EEG), we assessed cortical N1 responses evoked over fronto-midline regions (Cz) during balance recovery in response to backward support-surface perturbations loading both legs, as well as posterior-lateral directions that preferentially load the paretic or nonparetic leg. Cortical N1 responses were smaller and delayed in the stroke group. While older adults exhibited weak or absent relationships between cortical responses and clinical function, stroke survivors exhibited strong associations between slower N1 latencies and slower walking, lower clinical mobility, and lower balance function. We further assessed kinetics of balance recovery during perturbations using center of pressure rate of rise. During backward support-surface perturbations that loaded the legs bilaterally, balance recovery kinetics were not different between stroke and control groups and were not associated with cortical response latency. However, lateralized perturbations revealed slower kinetic reactions during paretic loading compared to controls, and to non-paretic loading within stroke participants. Individuals post stroke had similar nonparetic-loaded kinetic reactions to controls implicating that they effectively compensate for impaired paretic leg kinetics when relying on the non-paretic leg. In contrast, paretic-loaded balance recovery revealed time-synchronized associations between slower cortical responses and slower kinetic reactions only in the stroke group, potentially reflecting the limits of cortical engagement for balance recovery revealed within the behavioral context of paretic motor capacity. Overall, our results implicate individuals after stroke may be uniquely limited in their balance ability by the slowed speed of their cortical engagement, particularly under challenging balance conditions that rely on the paretic leg. We expect this neuromechanical insight will enable progress toward an individualized framework for the assessment and treatment of balance impairments based on the interaction between neuropathology and behavioral context.

## 1. Introduction

Despite our increasing knowledge of age-related shifts from primarily subcortically-to more cortically-mediated balance control, there is a limited understanding of how brain lesions, common in age-related diseases such as stroke, affect balance control. Slower motor reactions after stroke contribute to lower resilience to postural perturbations and increased fall risk.^1–4^ An impaired ability to rapidly and effectively use the paretic leg may require compensatory use of the nonparetic leg for whole-body behaviors such as balance and walking after stroke.^5–10^ From a neurophysiologic perspective, greater asymmetry in corticomotor excitability between paretic and nonparetic lower limbs, assessed in seated positions, is associated with greater reliance on the nonparetic leg to increase walking speed in individuals with chronic stroke.^11^ However, it is unclear this asymmetry in corticomotor neurophysiology, measured during seated tasks, translates to the control of whole-body movements. Recordings of brain activity during whole-body behaviors such as balance and walking may provide neuromechanical insight to help understand interactions between cortical activity and control of balance in post-stroke lower limb hemiparesis.

Lesions affecting cortical and subcortical pathways in older adults after stroke may compromise the ability to engage cortical resources for rapid balance recovery following destabilization. Using electroencephalography (EEG) during standing balance recovery reactions, we recently found that balance destabilization elicited greater cortical beta activity during balance recovery in neurologically-intact older adults who had relatively lower balance function than their peers^12^. This finding in older adults suggests greater sensorimotor cortical reliance for postural stability in individuals with lower balance function. Likewise, in neurotypical younger adults, cortical compensation during balance recovery may be reflected in larger cortical evoked responses during reactive balance in individuals with relatively poor balance ability^13^, when taking compensatory steps following challenging balance perturbations,^14,15^ and when perturbations are perceived as more threatening.^16,17^ Supporting this notion, individuals with lower post-stroke mobility commonly engage expansive cortical networks spanning sensorimotor and frontal regions during continuous walking tasks.^18^ In stroke, those with lower mobility function also reached a “ceiling effect” of lower cortical activity compared to higher-functioning individuals when presented with more challenging dual-task walking conditions.^18^ Together, these findings suggest individuals with stroke may increase reliance on cortically-mediated strategies for balance control, which may be compromised by lesions affecting cortical and subcortical structures.

Reactive balance control is essential to walking and mobility,^19^ but cortical engagement during the production of rapid corrective balance reactions to postural destabilization has not been characterized after stroke. Here, we measure the cortical N1 response, a large negative-going peak in the EEG signal over midline sensorimotor areas ∼150ms after a sudden disturbance to standing balance.^20^ The N1 response is thought to reflect detection of a sudden error to balance or posture.^21^ The N1 response has been localized to the supplementary motor area when constrained to a single source,^22,23^ but synchronization of multiple sources including the supplementary motor area, the anterior cingulate cortex, sensorimotor areas, and parietal cortex has been suggested to underlie the N1 response in time-frequency analyses.^24–26^ Neuromechanical investigation into cortical activity during balance reactions could also improve our understanding of temporal features of cortical engagement and relevance to behaviorally-relevant balance recovery responses necessary to prevent falls.

Delineating differences in cortical function involving paretic vs. nonparetic leg use during continuous mobility behaviors that involve bilateral use of lower limbs such as walking and balance is challenging. Individuals with lateralized cortical lesions due to stroke commonly present with limb hemiparesis, causing interlimb motor control deficits during balance and mobility behaviors.^7,19^ Differential cortical mechanisms during paretic and nonparetic leg motor activity have been identified during seated and isometric lower limb muscle contractions in individuals with chronic stroke.^11,27–29^ In particular, plantarflexor muscles^27^ play a key role in post-stroke mobility function^5,30–32^, and show more severely impaired corticomotor excitability (i.e. lower motor evoked potentials) compared to post-stroke corticomotor impairments across other lower limb muscle groups (e.g., dorsiflexors).^27^ Further, individuals with greater corticomotor excitability to plantarflexors in the nonparetic relative to the paretic leg show greater biomechanical reliance on the nonparetic leg to generate propulsive forces during walking, suggesting a link between corticomotor function and whole-body behaviors.^12^ Reactive balance paradigms may provide a method to assess neural contributions to whole-body behaviors through use of external perturbations that elicit a time-locked behavioral response that successively recruits subcortical followed by cortical contributions to lower limb motor reactions.^33–35^ Lateralized balance perturbations in stroke can mechanically load either the paretic or nonparetic leg during balance recovery, providing insight into lateralized deficits in balance recovery, in which individuals after stroke commonly sustain a fall.^1,10,19,36^ In a previous case series report by Solis-Escalante et al., direction-specific spectral components in evoked cortical N1 responses measured with EEG were present during reactive balance recovery in both older adults with stroke (n=3) and younger adult (n=6) participants.^37^ Specifically, lateralized perturbations elicited directional-specific spatial and spectral features within EEG recordings during the balance recovery response.^37^ However, whether these differences relate to clinical ability or balance impairment or potential differences in temporal versus spatial features of evoked cortical responses (e.g., timing and magnitude of response) during balance recovery in individuals post stroke has not been investigated. If changes in evoked cortical activity play a role in post-stroke balance impairment and increased fall risk, a better understanding of this role could help identify new therapeutic targets to reduce fall risk after stroke.

Our central hypothesis is that stroke impairs the ability to rapidly and effectively engage the cerebral cortex in balance-correcting behavior, resulting in asymmetrical interlimb contributions to post-stroke mobility behavior. In the present study, we used multidirectional standing balance perturbations to differentially challenge balance control between limbs. We assessed kinetic reactions and the speed and magnitude of cortical engagement during balance recovery in individuals with and without post-stroke lower limb hemiparesis. We further tested the effect of mechanical balance perturbations loading either the paretic or nonparetic leg during balance recovery on evoked cortical N1 responses and kinetic reactions and relationships to clinical balance and mobility function. We predicted that 1) stroke survivors would have later and attenuated perturbation-evoked cortical N1 responses during balance recovery compared to neurotypical, age-matched controls, with the most impaired cortical N1 responses during paretic-loading conditions and 2) that longer latencies of perturbation-evoked cortical N1 responses would be associated with clinical balance deficits and slower kinetic reactions after stroke.

## 2. Materials and methods

### 2.1. Study design and participants

Eighteen individuals with chronic (>6 mo.) stroke (**Table 1**) and 17 age-matched controls were recruited. Inclusion criteria included above the age of 21, the ability to walk at least 10 meters without the assistance of another person, the ability to stand unassisted for at least 3 minutes, and the cognitive ability for informed consent. Participants were excluded for any diagnosed neurologic condition other than stroke or pain affecting standing or walking. The experimental protocol was approved by the Emory University Institutional Review Board and all participants provided written informed consent.

**Table 1.** Stroke participant characteristics (n=18)

| Age (yrs) | Gender (M/F) | PSD (months) | Paretic side | Lesion Location | Gait speed (m/s) | TUG (s) | miniBEST |
| --- | --- | --- | --- | --- | --- | --- | --- |
| 65-69 | M | 122 | R leg | L BG | 1.25 | 8.61 | 23 |
| 75-79 | F | 43 | L leg | R IC/BG | 0.99 | 12.42 | 21 |
| 60-64 | M | 56 | R leg | L IC/BG | 0.33 | 37.00 | 11 |
| 65-69 | F | 24 | L leg | L Striatum, IC, Caudate | 1.14 | 9.73 | 23 |
| 55-59 | F | 14 | L leg | N/A | 0.45 | 20.94 | 14 |
| 65-69 | F | 128 | L leg | R CR, M1, S1 | 0.70 | 12.96 | 25 |
| 80-84 | M | 89 | L leg | R MCA | 0.70 | 19.59 | 12 |
| 45-49 | F | 41 | L leg | R PLIC | 0.80 | 11.70 | 22 |
| 65-69 | F | 85 | R leg | L IC | 1.06 | 11.83 | 19 |
| 55-59 | M | 56 | L leg | R ACA | 0.37 | 35.55 | 2 |
| 75-79 | M | 40 | R leg | L Pons | 0.52 | 22.55 | 14 |
| 55-59 | M | 86 | L leg | R frontal, parietal | 0.64 | 20.04 | 11 |
| 90-94 | M | 6 | L leg | N/A | 0.48 | 24.55 | 13 |
| 40-44 | F | 36 | R leg | L MCA | 0.83 | 11.56 | 20 |
| 55-59 | M | 49 | L leg | N/A | 1.36 | 8.93 | 21 |
| 50-54 | M | 48 | L leg | R PLIC | 1.00 | 21.53 | 15 |
| 70-74 | F | 114 | L leg | R ACA | 0.55 | 28.19 | 10 |
| 65-69 | F | 83 | R leg | L PLIC, striatum | 0.53 | 21.60 | 11 |

PSD = post stroke duration; TUG=Timed-Up-and-Go; m/s=meters per second; M1: primary motor cortex; S1: primary somatosensory cortex; IC: internal capsule; BG: basal ganglia; ACA: anterior cerebral artery; MCA: middle cerebral artery; PLIC: posterior limb IC; CR: corona radiata N/A = not available.

Participants completed a single visit of clinical balance and mobility testing (i.e., miniBEST,^38^ Timed-Up-and-Go (TUG),^39^ 10-meter walk test) following standard clinical practice procedures and administered by the same licensed physical therapist. Participants were then subjected to a series of support-surface translational perturbations to assess EEG measures of evoked cortical activity and biomechanical reactions during standing balance recovery.

### 2.2. Standing balance perturbations

Participants stood barefoot on a moving platform (Factory Automation Systems, Atlanta, GA) and were subjected to anterior, posterior, and left-ward and right-ward posterolateral support-surface translational perturbations that served to preferentially load either the paretic leg, the nonparetic leg, or equal legs during balance recovery. During the paretic-loaded condition, the support-surface moves posterolaterally towards the nonparetic leg, shifting a greater proportion of body weight support onto the paretic leg (**Figure 1**). Likewise, the nonparetic-loaded condition shifts a greater proportion of body weight support onto the nonparetic leg. In contrast, in the bilateral condition, the support-surface moves in the posterior direction with no lateralization (**Figure 1**), targeting plantarflexor agonist muscles to correct for postural destabilization. Anteriorly directed perturbations were also included to prevent participants from leaning backward in anticipation of posteriorly directed perturbations used in theanalyses. Participants were instructed to stand with their typical, self-selected posture and foot placement, as similar motor response latencies are observed across a range of narrow and wider stances.^40^ Twenty-four perturbations (7.5 cm, 16.0 cm/s, 0.12 g) within each of the four directions (total of 96 perturbations) were delivered in a pseudorandomized order at unpredictable inter-trial intervals (15 – 60s). Participants received instructions to recover balance with a feet-in-place strategy if possible and to keep arms folded at their chest. We selected this relatively low-level perturbation level because it could be successfully completed by most participants using a feet-in-place strategy. However, two participants in the stroke group and one control participant were unable to recover balance at this perturbation level with feet-in-place; for these participants the magnitude of balance perturbation was scaled down to (6.0 cm, 12.0 cm/s, 0.08 g) while maintaining the same temporal characteristics of the perturbation. The perturbation series was delivered in a pseudorandomized order, where a perturbation of the same direction had no more than 2 consecutive occurrences. Real-time EEG activity and ground reaction force levels were monitored by the experimenter to ensure that the participant returned to baseline body position and levels of cortical and muscle activity after each trial before the next perturbation was delivered. Instances of stepping were noted in real-time for offline confirmation and exclusion from analyses based on ground reaction forces.^15^ Participants took a seated rest break every 8 minutes during balance perturbation testing, or more frequently if the participant requested a break or reported or showed signs of fatigue during testing.

**Figure 1:**
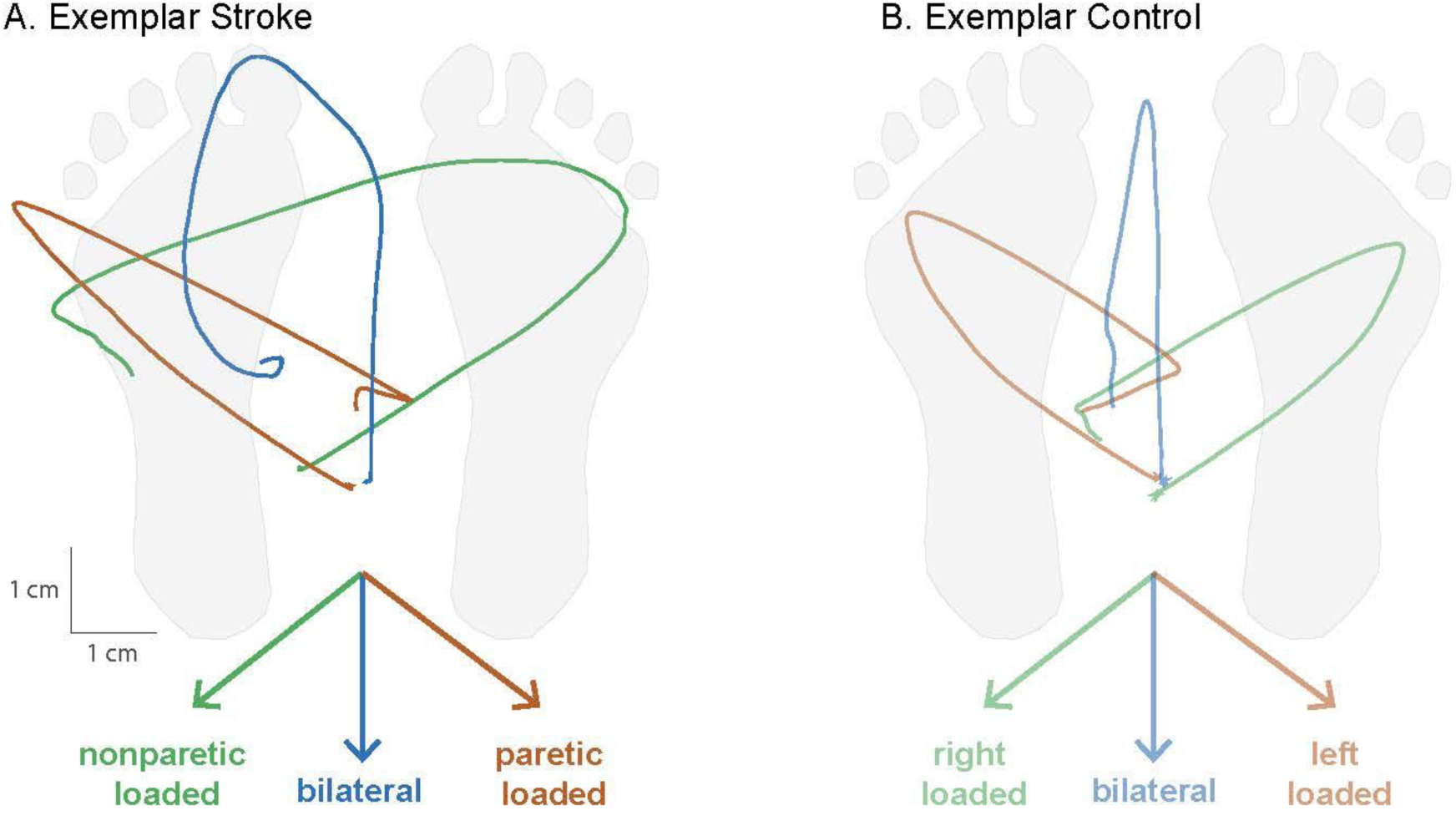
Perturbation conditions and resulting kinetic reactions for balance recovery. Across each of the perturbation conditions, the support surface moved in the direction indicated by the colored arrows, displacing the center of mass towards the paretic leg, bilateral legs, or nonparetic leg (**A**), necessitating rapid corrective shifts in center of pressure to prevent imbalance. Mechanical effects of each condition on center of pressure displacement trajectories (condition-averaged across trials) and postural loading are depicted in an exemplar stroke (**A**) and control (**B**) participant. In lateralized conditions, the center of pressure trajectory was shifted towards the paretic or nonparetic legs (green and orange), while the bilateral condition showed no lateralized bias (blue). Note that the paretic-loaded condition refers to movement of the support surface and feet in the direction of the nonparetic limb, consequently shifting the center of pressure beneath of paretic limb and loading the paretic limb during the rapid kinetic reaction. Condition-averaged center of pressure trajectories are shown from an example participant in each group.

### 2.3. EEG data acquisition and analyses

During balance perturbations, cortical activity was continuously recorded from EEG signals using a 64-channel active electrode cap (actiCAP, actiCHamp amplifier, Brain Products, GmbH, Gilching, Germany). EEG signals were digitized with a 24-bit analog-to-digital converter and an online 20 kHz low-pass filter and before sampling at 1000 Hz and storing for offline analysis. All EEG data were preprocessed using freely available functions from the EEGlab toolbox and custom MATLAB scripts.^41^ Continuous data time-locked to the perturbation onset were imported into EEGlab. Trigger labels for successful feet-in-place trials (i.e., no reactive step taken) were selected across all conditions. Continuous EEG data were high-pass filtered (cutoff 0.5 Hz, finite impulse response, filter order 3300) and downsampled to 500 Hz. Bad channels were identified through visual inspection, then removed and interpolated. Data were re-referenced to an average reference. Line noise was removed using the Cleanline plugin.^41^ Data were then epoched -2 to 2 seconds around each perturbation (platform onset at t=0 s), and decomposed into maximally independent components (ICs) using adaptive mixture component analysis algorithm (AMICA).^42^ ICs from AMICA were categorized using the ICLabel plugin, an automated algorithm that identifies nonbrain sources (e.g., eye, muscle, and cardiac activity) and brain sources,^43^ and confirmed with visual inspection.

Nonbrain sources were removed. The remaining brain ICs were projected back into channel space. For one participant with a lower number of non-stepping trials, we aimed to maximize the number of trials for AMICA by including all trials (step and no step) in this part of the preprocessing pipeline before removing all stepping trials for N1 waveform computation and all subsequent analyses. Data were visually inspected and trials with excessive signal drift were removed.

EEG data from the midline sensorimotor region (Cz) for posterior and postero-laterally directed perturbations were selected and low-pass filtered at 30Hz for evoked cortical event-related potential analyses and baseline subtracted (-150 to -50ms).^20,44^ The peak latency and amplitude of thecortical N1 response were extracted from the mean waveform across all trials as well as the mean waveform across each condition. The cortical N1 response was defined as the first local minimum point of negative value in the EEG waveform within 100-300ms post-perturbation. As individuals with stroke tended to show polyphasic perturbation-evoked cortical responses (**Figure 3C**), this automated selection criteria enabled consistency in selection of the cortical N1 response within-participants (between conditions) and between-participants. Two participants required an extended time window of 100-350ms post-perturbation because the first local minimum in the cortical response waveform occurred > 300ms post-perturbation (316 and 314ms post-perturbation for all conditions collapsed, respectively)

### 2.4. Kinetic data acquisition and analysis

Kinetic (AMTI OR6-6 force plates) and kinematic (10-camera Vicon Nexus 3D motion analysis system) data were recorded during balance perturbations (100 Hz sampling frequency). Reflective markers were placed on anatomical landmarks on the legs and trunk (e.g., head, neck, hips, knees, ankles, feet) and were used as inputs to Vicon’s plug-in-gait model to compute the body’s center of mass velocity and displacement throughout balance recovery.

The corrective kinetic reaction during balance recovery following perturbations was quantified as the center of pressure (CoP) rate of rise (RoR). During the perturbation, the CoP initially moves passively as a result of the perturbation, and the individual must then rapidly counteract this effect to slow and reverse the direction of CoP movement to maintain upright stability (**Figure 1**).^45^ The CoP RoR in the later 150-300ms phase of balance recovery occurs during a timeframe in which an individual’s active contribution to reactive balance is possible and necessary for a successful feet-in-place balance recovery. As such, the average slope of this later phase CoP position trajectory indexes how quickly an individual generates corrective responses to the loss of balance. A slower CoP RoR relative to the support surface movement would result in a less effective neuromechanical stabilization strategy that may lead to loss of balance.^45^ The CoP position was used to assess CoP RoR in the direction parallel to support-surface movement quantified as the linear slope (i.e., rate of change in CoP position) between 150-300 ms post-perturbation onset.

### 2.5. Statistical analyses

We confirmed normality and heterogeneity of variance of all data used for analyses using Kolmogorov-Smirnov and Levene’s tests, respectively. We matched lateralized balance conditions of paretic-loading to left-loading and nonparetic-loading to right-loading in controls. First, we compared cortical N1 response latency and amplitudes collapsed across all conditions between stroke and control groups using independent t-tests. We then tested group (control, stroke) and condition (bilateral, paretic-loaded, nonparetic-loaded) main effects and interactions between group and condition for each cortical N1 metric (peak latency, peak amplitude) using a two-way analysis of variance (ANOVA). We used Pearson product moment correlation coefficients to test for associations between cortical N1 peak metrics and clinical and biomechanical metrics. We used multiple linear regression (MLR) analyses (factors: group, N1, group-by-N1) to test whether the relationship between cortical N1 peak metrics (latency or amplitude) and clinical metrics (walking speed, TUG, or miniBEST) differed as a function of group in the collapsed N1 waveform and across each condition. We used a two-way ANOVA (factors: group, condition, group-by-condition) to test for group-by-condition interaction and main effects on CoP RoR. We used MLR analyses (factors: group, N1, group-by-N1) to test whether the relationship between cortical N1 peak metrics (latency or amplitude) and CoP RoR differed as a function of group across each condition. All analyses were performed using SPSS version 27 with an a-priori level of significance set to 0.05.

## 3. Results

One participant in the control group withdrew from the study due to fear of falling; this participant was unable to complete balance perturbation testing and was excluded from analysis. Due to fatigue and increased time necessary for balance testing, eight participants in the stroke group completed a shortened protocol (range of perturbations completed: 60-86 out of 96 perturbations in full protocol). Including those adopting the shortened protocol, EEG recordings from18 individuals post-stroke and 16 controls were included in analyses. Participants in the stroke group had lower clinical measures of balance function on the miniBEST (p<0.001), slower performance on the TUG-test (p<0.001) and slower walking speed (p<0.001) (**Table 2**). Technical issues involving biomechanical force data acquisition occurred in three participants (stroke, n=2; control, n=1); these participants were excluded from analyses involving CoP RoR, but are included in all other analyses.

**Table 2.** Participant group characteristics.

|  | Stroke, n=18 | Older adult controls, n=16<sup>a</sup> | P-value (t-test) |
| --- | --- | --- | --- |
| Age, years | 65 ± 12 | 69 ± 8 | 0.241 |
| Gender, male/female | 9/9 | 4/12 | 0.126<sup>b</sup> |
| miniBEST score | 16 ± 6 [2-25] | 24 ± 2 [20-28] | <0.001 |
| TUG-test, seconds | 18.8 ± 8.7 [8.6-37.0] | 8.8 ± 1.9 [5.7 – 11.8] | <0.001 |
| Walking speed, m/s | 0.76 ± 0.31 [0.33 – 1.36] | 1.22 ± 0.16 [1.02 – 1.56] | <0.001 |

TUG=Timed-Up-and-Go; m/s=meters per second; <sup>a</sup>Excludes control participant (n=1) who withdrew from the study and was not included in analyses. Values are depicted in mean ± standard deviation [range: minimum - maximum]. <sup>b</sup> Fisher's exact test

### 3.1. Effect of stroke on cortical N1 response and relationship to clinical balance and mobility

Individuals post stroke exhibited delayed latencies to the cortical N1 peak (stroke = 219 ± 39 ms; control = 196 ± 22 ms, *p* = 0.025) and reduced N1 amplitudes (stroke = 14.9 ± 11.9 µV; control = 21.7 ± 11.0 µV, p = 0.047) compared to age-matched controls across all conditions (**Figure 2A**). The relationship between N1 latencies and behavioral outcomes varied between groups. In the stroke group, delayed N1s correlated with lower miniBEST scores (r = -0.61, p = 0.007), slower Timed-Up-and Go-(TUG) test performance (r = 0.53, p = 0.024), and exhibited a trend with reduced walking speed (r = -0.46, *p* = 0.055) (**Figure 2B**). In the control group, delayed N1s were similarly associated with slower TUG test performance (r = 0.508, *p* = 0.045) and showed no correlation with miniBEST scores (r = -0.274, p = 0.304) or walking speed (r = -0.001, p = 0.997). Examining N1 response latency in each condition separately showed similar relationships across all conditions within the stroke and control groups (not shown). While the stroke group consistently showed stronger relationships between cortical N1 latency and clinical metrics compared to controls (**Figure 2B)**, group-by-N1 latency interactions failed to meet our *a priori* level of significance across clinical metrics (miniBEST (*t*= -1.25, *p*=0.220), TUG test performance (*t*= 1.00, *p*=0.325), walking speed (*t*=-1.10, *p*=0.279). There were no associations between N1 amplitude and any clinical metric in the stroke or control groups (all *p*>0.11). There were no significant group-by-N1 amplitude interactions for any clinical metric (miniBEST, *p*=0.333; TUG, *p*=0.906; gait speed, *p*=0.979) (**Figure 2B).**

**Figure 2:**
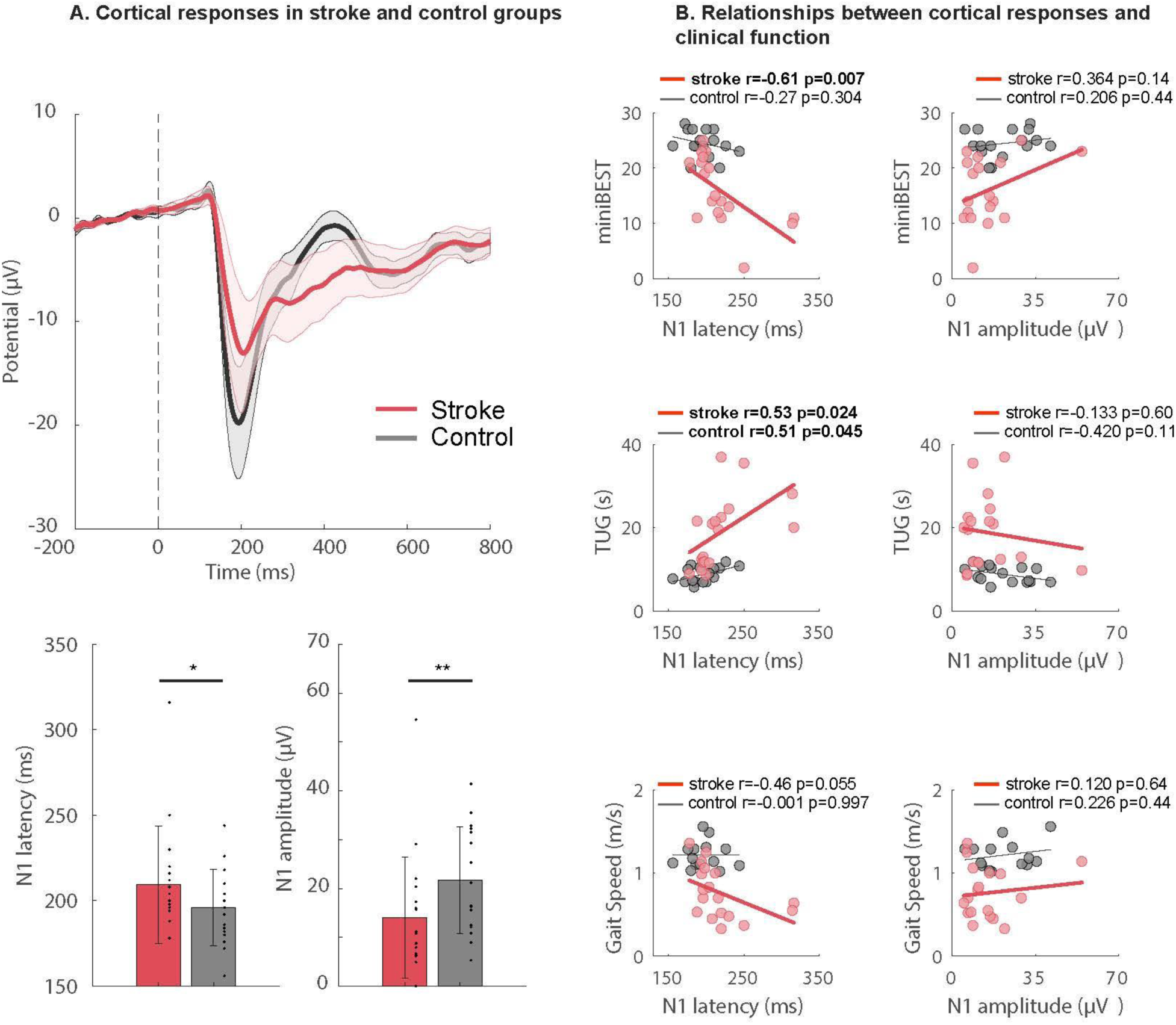
Impaired cortical responses associated with clinical balance and mobility after stroke. **(A)** Evoked cortical N1 response waveforms during balance recovery are shown collapsed across all perturbation conditions in each group. Means ± SEMs are depicted in the shaded regions (top). In participants after stroke, cortical N1 responses showed later peak latencies (stroke=219±39 ms; control=196±22 ms, \**p*=0.025) and attenuated peak amplitudes (stroke=14.9±11.9; control=21.7±11.0, *\*\*p*=0.047) compared to age-matched controls (bottom). **(B)** Relationships between evoked cortical N1 peak latency (left column) and peak amplitude (right column) during reactive balance across all conditions versus miniBEST score (top row), single-task timed-up-and-go (TUG) test (middle row), and walking speed (bottom row), in individuals post stroke and age-matched controls. For the miniBEST, slower cortical N1 response latencies were associated with lower miniBEST score in the stroke group (r=-0.61, *p*=0.007) while showing no relationship in controls (r=-0.274, p=0.304). **(A)**. For the TUG, both groups showed an association between later cortical N1 latency and slower TUG performance (stroke: (r=0.53, *p*=0.024; controls: r=0.508, *p*=0.045). For gait speed, slower cortical N1 response latencies showed a trend for association with slower gait speed in the stroke group (r=-0.46, *p*=0.055), a relationship that was absent in controls (r=-0.001, *p*=0.997). Cortical N1 peak amplitude (right column) was not significantly associated with any clinical balance or mobility function metric in the stroke and control groups (*p*>0.11).

### 3.2. Effect of Lateralization of Balance Perturbations in Stroke and Controls

Contrary to our initial hypotheses, the lateralization of balance perturbations did not exhibit any discernible impact on cortical N1 response latency or amplitude in either the stroke or control groups (**Figure 3A, B**). Despite considerable between-individual variability (Figure 3C), no significant interaction effects were observed for N1 latency (*F*_2,64_=0.463, *p*=0.722) or amplitude (*F*_2,64_=0.624, *p*=0.516) (**Figure 3B**).

**Figure 3:**
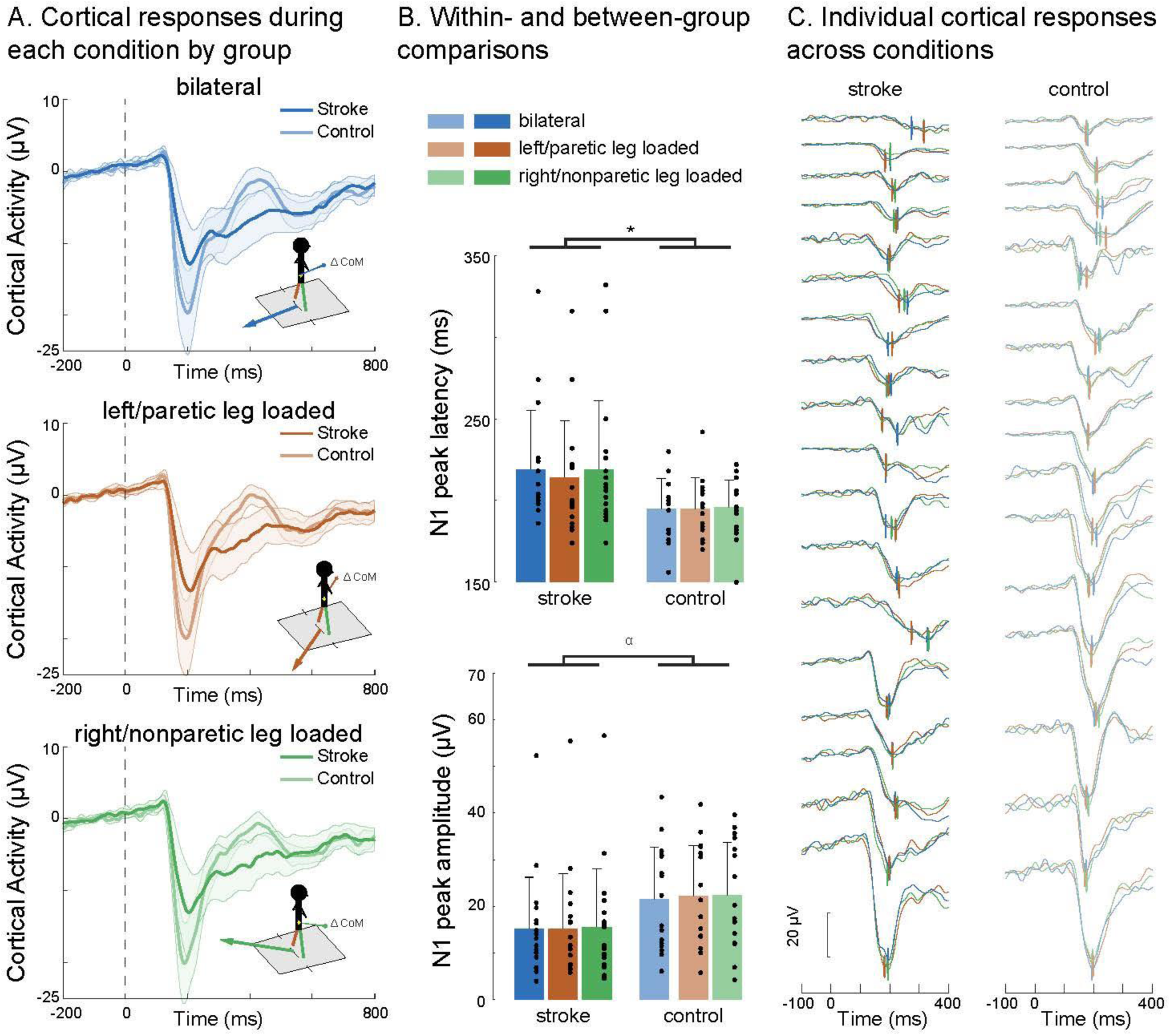
Cortical N1 responses within each balance perturbation conditions. **(A)** Cortical N1 response waveforms evoked during symmetrical bilateral (top) and lateralized perturbation conditions loading each the left/paretic (middle) and right/nonparetic legs (bottom) are shown in each the stroke and control groups. Means ± SEMs are depicted in the shaded regions. **(B)** There was a main effect of group, in which stroke showed later N1 peak latencies (*F*_1,32_=5.27, \**p*=0.028) (top) and a trend towards lower N1 peak amplitudes (*F*_1,32_=2.932, α *p*=0.097) (bottom) compared to controls. While the paretic-loaded condition tended to elicit faster N1 response latencies (214±35 ms) compared to the nonparetic-loaded (219±42 ms) or bilateral condition (219±42 ms) within the stroke group, among high between-participant variability **(C)** there were no main effects of condition for N1 peak latency (*F*_1,32_=1.063, *p*=0.310) (top) or amplitude (*F*_1,32_=1.47, *p*=0.292) (bottom). Means ± SDs depicted in figure bar plots.

Interestingly, in the stroke group, the paretic-loaded condition tended to evoke faster N1 response latencies (214 ± 35 ms) compared to the nonparetic-loaded (219 ± 42 ms) or bilateral condition (219 ± 42 ms) (**Table 3**) (**Figure 3C**). However, statistical analyses for N1 peak latency revealed significant main effects of group (*F*_1,32_=5.27, *p*=0.028) but not for condition (*F*_1,32_=1.063, *p*=0.310). Regarding N1 amplitude, a non-significant trend toward a main effect of group (*F*_1,32_=2.932, *p*=0.097) was observed, with no main effects of condition (F _1,32_=1.47, *p*=0.292) (**Figure 3B**).

**Table 3.** Group and condition differences in cortical and kinetic responses.

|  | Stroke, n=18 | Older adult controls, n=16<sup>a</sup> | P-value |
| --- | --- | --- | --- |
| **Cortical N1 response latency (ms)** |  |  | 0.722 (interaction) |
| Collapsed | 219±39 | 196±22 | 0.025 (between-group t-test) |
| Bilateral leg loaded | 219±36* | 195±19* | 0.310 (condition main effect) |
| Paretic leg loaded | 214±35* | 195±19* | NA |
| Nonparetic leg loaded | 219±42* | 194±20* | NA |
| **Cortical N1 response amplitude (µV)** |  |  | 0.516 (interaction) |
| Collapsed | 14.9±11.9 | 21.7±11.0 | 0.047 (between-group t-test) |
| Bilateral leg loaded | 15.1±11.1<sup>∞</sup> | 21.4±11.2<sup>∞</sup> | 0.292 (condition main effect) |
| Paretic leg loaded | 15.1±12.0<sup>∞</sup> | 22.3±10.8<sup>∞</sup> |  |
| Nonparetic leg loaded | 15.5±12.5<sup>∞</sup> | 22.3±11.4<sup>∞</sup> |  |
| **Kinetic CoP Rate of Rise (cm/s)** |  |  | **0.026** (interaction) |
| Bilateral leg loaded | 28.8±7.8 | 31.6±4.6 | 0.312 (vs. P loaded) |
| Paretic leg loaded | 27.6±9.3** | 35.2±5.9** |  |
| Nonparetic leg loaded | 35.2±12.2 | 35.7±5.7 | **0.004** (vs P loaded),<br>**0.002** (vs. bilateral) |

Main effect of group indicated by \* at p=0.028 and <sup>∞</sup> at p=0.097. Between-group post-hoc testing \*\*p=0.012; mean ± standard deviation.

### 3.3. Kinetic reactions during balance recovery and associations with cortical N1 response

There was a group-by-condition interaction effect on the center of pressure (CoP) rate of rise (RoR) (F_2,29_=3.054, p=0.026), in which the stroke group had a slower CoP RoR than the control group only within the paretic-loaded condition (*p*=0.012) (**Figure 4A&B**). Within the stroke group, the CoP RoR was faster in the nonparetic-loaded condition compared to each the paretic-loaded condition (*p*=0.004) and the bilateral condition (*p*=0.002), with no difference between the paretic-loaded and bilateral conditions (*p*=0.312). Within the control group, the CoP RoR was slower in the bilateral condition compared to each the lateral loading conditions (left-loaded, *p*<0.001; right-loaded, *p*=0.012). The lateral loading conditions in controls were not different from each other (*p*=0.312) Within the stroke group, later N1 peak latencies were associated with slower CoP RoR within the paretic-loaded condition (*r*=-0.70, *p*=0.003), but not in the nonparetic-loaded condition (*r*=-0.41, *p*=0.11) or the bilateral condition (*r*=-0.27, *p*=0.313) (**Figure 4B**). None of these associations were observed in the control group (all *p*>0.138). When testing for group-by-N1 latency interaction effects on CoP RoR, there was a trend for interaction within the paretic-loaded condition (*t*= -1.803, *p*=0.083) but not the nonparetic-loaded (*t*= -0.012, *p*=0.991) or bilateral conditions (*t*= -0.705, *p*=0.487). For N1 amplitude, there were no group-by-N1 amplitude interaction effects in any balance condition (all *p*>0.598) or relationships with CoP RoR for any condition in either group (all *p*>0.491).

**Figure 4.**
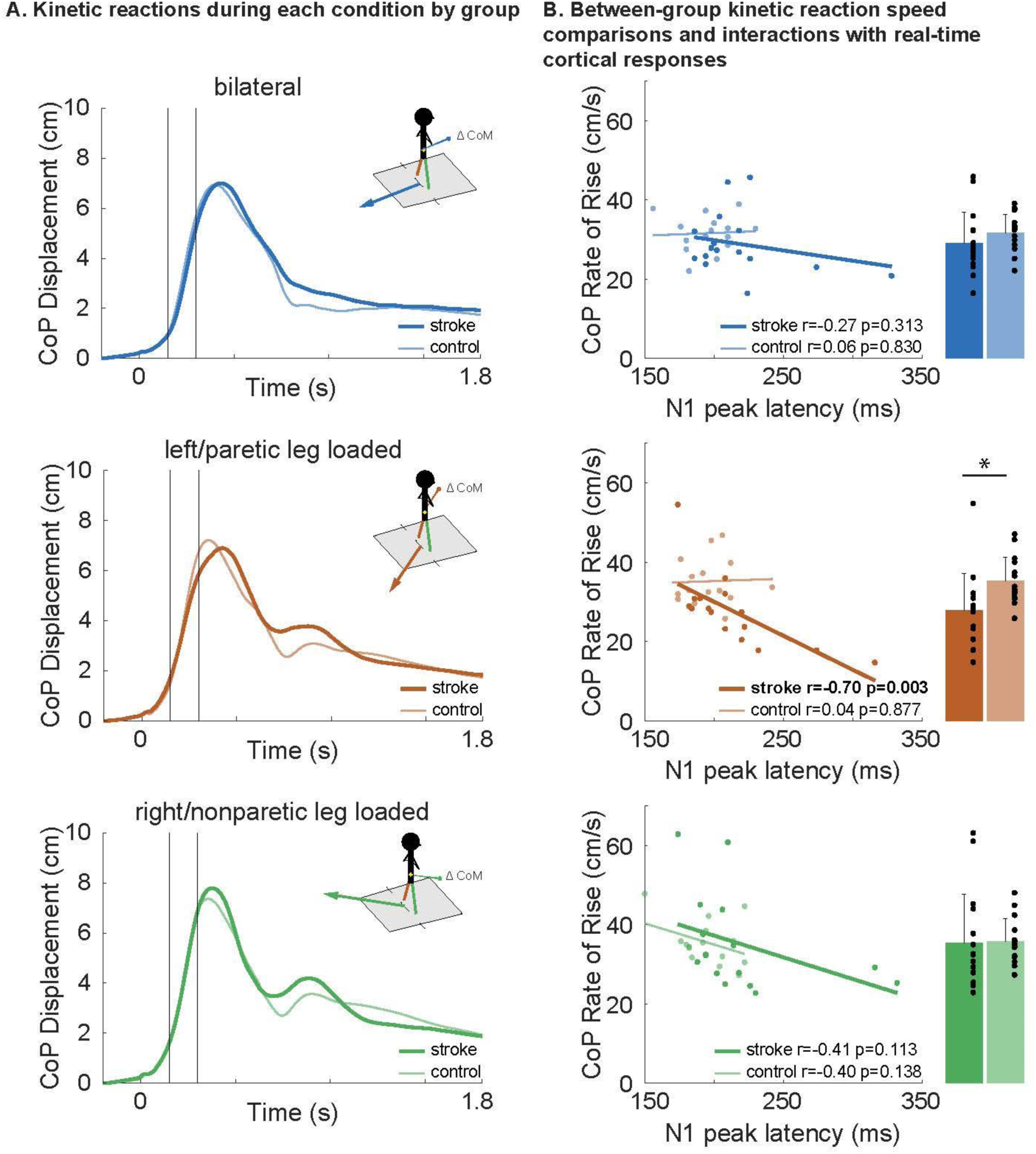
Balance recovery kinetics and relationship to time-synchronized cortical N1 response latency. **(A)** Center of pressure displacement across bilateral, nonparetic-loaded, and paretic-loaded perturbation conditions are shown as mean waveforms for each group. The center of pressure rate of rise (CoP RoR) was calculated as the linear slope of the CoP displacement between 150-300 ms (black vertical lines) post-perturbation onset (time=0). During paretic-loaded balance recovery (middle), participants with stroke showed slower CoP RoR compared to nonparetic-loaded recovery (bottom) (*p*=0.004) and controls (*p*=0.012) and no difference compared to the bilateral condition (top). During nonparetic-loaded balance recovery, there was no difference in CoP RoR between groups (bottom). Mean ± SD are shown for CoP RoR. **(B)** During paretic-loaded balance recovery (middle), there was a relationship between later cortical N1 peak latencies and slower CoP RoR in stroke (r=-0.70, *p*=0.003) while no effect was observed in controls. During nonparetic-loaded recovery (bottom) and bilateral conditions (top), there was no relationship between cortical N1 latency and CoP RoR. No relationships were observed between N1 amplitude and CoP RoR in any condition or group (not shown).

## 4. Discussion

The observed findings are the first to demonstrate that clinical and kinetic balance dysfunction in people with post-stroke lower limb hemiparesis is related to delayed cortical N1 responses evoked during reactive balance recovery. Our reactive balance paradigm provided a well-controlled probe of cortical reactivity during a functionally-relevant, whole-body behavior, demonstrating that balance perturbations elicit slower, smaller cortical responses after stroke compared to age-similar controls. These findings are consistent with the notion of generally impaired cortical engagement for balance control in people after stroke and may reflect altered cortical mechanisms underlying balance and gait dysfunction. In particular, a reduced ability to rapidly engage cortical resources during balance recovery may contribute to balance and mobility dysfunction post-stroke, supported by relationships between slower N1 responses and slower mobility/lower balance function that were present only in the stroke group. Further, while balance conditions loading the paretic leg resulted in slower kinetic reactions for balance recovery compared to controls,^2–4^ balance conditions that positioned the nonparetic leg for recovery enabled individuals post stroke to achieve similar kinetic reactions to their age-matched peers. Relationships between time-synchronized cortical response speed and kinetic reactivity during paretic-loaded balance recovery may reflect the constraints of rapid cortical engagement at the limits of paretic motor capacity (e.g., paretic leg loading) that are masked when the nonparetic leg is engaged in compensatory balance control (i.e., nonparetic leg loading and bilateral loading conditions). Together, our findings suggest that temporal features of evoked cortical N1 responses during reactive balance recovery may provide a useful biomarker of clinically-relevant balance and mobility behavior that may serve as a target for rehabilitation efforts aimed at maximizing independence and reducing fall risk in the chronic stage of stroke recovery.

### 4.1. Impaired cortical engagement may contribute to mobility deficits post stroke

One of the most consistent neurophysiologic findings post-stroke is slowed and reduced cortical excitability within the lesioned primary motor cortex^46^ that may explain, in part, the slower and lower magnitudes of evoked cortical N1 peak responses compared to controls. As such, stroke-related effects in older adults may compromise the typical engagement of cortical resources in the aging brain for balance control.^13,20,47^ Longer latencies and smaller amplitudes of peak cortical N1 responses in people with stroke (**Figure 2A**) are consistent with the presence of impaired cortical engagement in balance recovery, and were driven by individuals with the most impaired balance and mobility function (**Figure 2B**). Impaired cortical engagement may be particularly detrimental during abrupt and challenging balance perturbations that elicit greater corticomotor drive to for balance recovery compared to less abrupt balance perturbations, as evidenced in neurotypical individuals by increases in functional connectivity between cortical activity and reactive lower limb motor responses with more challenging perturbations.^48^ Together, these findings suggest that the speed and effectiveness of sensorimotor error detection and information processing is compromised in individuals with cortical and subcortical lesions.

The presence of brain-behavioral relationships only in the stroke group may indicate a greater need to rapidly detect balance errors (i.e., reflected in the cortical N1 response latency)^21^ for motor control influencing balance and mobility behaviors after stroke. The ability for rapid error detection, potentially reflected in the N1 response, may play a distinct role from that of other information encoded within the cortex. Cortical error detection speed (i.e., reflected in the cortical N1 latency) may be an aspect of balance control that limits stroke balance ability, but may not be the limiting factor in neurotypical older adults due to a wider range of heterogeneous factors (i.e., balance confidence, cognitive flexibility, attention ability, greater automaticity of balance control) that may contribute to brain-balance relationships.^20^ Similarly, we recently observed relationships between measures of N1 timing and amplitude and measures of balance and mobility in a group of individuals with Parkinson’s disease that were not present in the control group.^44^ The presence of brain-behavioral relationships when collapsing data across all direction conditions in the stroke group (**Figure 2B**) may reflect the bilateral leg performance necessary during post-stroke balance and walking behavior assessed in clinical contexts. These relationships further suggest that the inability to engage the cortex rapidly and effectively for balance control may limit potential recovery of clinical balance and mobility function after stroke, as illustrated in the most severely impaired individuals after stroke (**Figure 2B)**. The high within-group variability in cortical responses and clinical metrics is consistent with high variability in balance and walking function after stroke.^49–51^ Together, the present findings reveal neurophysiologic features of cortical slowness that are linked to balance and mobility dysfunction after stroke, potentially contributing to increased falls risk in individuals post stroke.^3^

### 4.2. Lateralization of balance perturbations did not affect cortical responses

Similar cortical N1 response latencies and amplitudes elicited during (more impaired) paretic versus (less impaired) nonparetic-loaded balance recovery conditions may reflect different neuromechanical features of balance recovery after stroke. While the lateralization of perturbations towards paretic and nonparetic legs generated asymmetrical limb loading and balance recovery (**Figure 1**), it was surprising that perturbation loading condition did not affect cortical responses within the stroke group (**Figure 3A-B**). While nonparetic-loaded cortical N1 response speeds may reflect relatively faster sensorimotor integration and motor reactivity of the nonparetic leg (**Figure 4**), paretic-loaded cortical N1 response speeds may reflect heightened surprise, threat, and/or error detection^17,21^ that occurs with increased loading towards the more undesirable leg for weight bearing and motor control.^7^ The latter may explain the (non-significant) tendency for individuals post-stroke to show faster cortical N1 response latencies during paretic-loaded conditions (**Figure 3B)**.

Nonetheless, one previous study found preliminary evidence (n=3) for direction-specific effects of balance perturbations on spectral features of evoked cortical N1 responses after stroke,^37^ supporting the possibility that directional information may be encoded in spatial and spectral features of EEG recordings during balance recovery not assessed in the present study and others reporting no directional effect.^52^ Together these findings illustrate the behavioral relevance of temporal features of evoked cortical activity and motivate future studies investigating event-related spatial and spectral features, which may identify potential subgroups within a larger cohort of people with post-stroke lower limb hemiparesis during lateralized balance recovery.

### 4.3. Impaired and compensatory post-stroke kinetic reactions occur during lateralized balance recovery

The present findings provide evidence that people post stroke can achieve similar kinetic reactive balance performance to their age-matched peers when they are mechanically positioned to compensate with the nonparetic leg. While the stroke group demonstrated slower kinetic balance reactions during paretic loading compared to controls (**Figure 4**), they showed faster and comparable kinetic balance reactions to their age-matched peers during nonparetic loading. This finding during reactive balance builds upon previous research showing some individuals post-stroke demonstrate slower paretic leg balance reactions,^2–4^ yet are able to effectively compensate for severe paretic leg impairment through increased nonparetic leg postural reliance, with a shift towards more lateralized balance control with the nonparetic leg.^7^ Slower kinetic CoP RoR responses during bilateral leg recovery compared to the lateralized conditions within the control group may reflect different biomechanical conditions presented by medial-lateral compared to anterior-posterior balance perturbations, which may differ in difficulty.^53^ Notably, the slower kinetic reactions in bilateral compared to lateralized perturbations within the control group was in contrast to the stroke group, which demonstrated comparable kinetic reactions in the bilateral and lateralized paretic-loaded condition (**Figure 4A**). Thus, it is possible that controlling for biomechanical differences and balance challenge presented by medial-lateral and anterior-posterior directional postural perturbations (e.g., adopting tandem stance in baseline standing posture) could reveal the effect of paretic leg loading on kinetic reactions for balance recovery after stroke in anterior-posterior conditions. The present findings provide a foundation for future rehabilitation studies to test whether therapeutic strategies aimed at accelerating nonparetic leg balance reactions could effectively improve the post-stroke balance recovery ability, particularly in individuals with limited recovery potential of the paretic lower limb.

### 4.4. Paretic-loaded balance recovery reveals cortical-kinetic interactions after stroke

Regardless of the neural origin, the present results suggest that faster cortical engagement in response to balance perturbations is linked to more rapid speed of the subsequent kinetic reactions during balance recovery. While there was not an effect of balance condition on cortical N1 responses or relationships to clinical ability, paretic-loaded balance recovery revealed time-synchronized relationships between the speed of cortical N1 responses and the speed of corrective kinetic reactions (**Figure 4**), potentially linked to lower resilience to postural destabilization.^19,33^ Cortical-kinetic relationships were absent in controls and in bilateral and nonparetic-loaded conditions in stroke, suggesting that positioning the paretic leg for balance recovery unmasks the cortical limits of balance recovery after stroke. Future studies are needed to test whether manipulation of threat or somatosensory information differentially influence the speed of evoked cortical N1 responses and its effect on time-synchronized kinetic reactions when biasing paretic or nonparetic legs for balance recovery.

Different relationships in time-synchronized cortical-kinetic responses across balance conditions further suggest that post-stroke lower limb hemiparesis may drive individuals to engage different neural strategies for balance recovery compared to controls. While not statistically significant, a similar relationship between cortical N1 responses and kinetic reactions during the nonparetic-loaded condition is interesting because it suggests that individuals post stroke may achieve similar reactive balance performance through more cortically-mediated balance responses to their age-matched counterparts, potentially with compensatory use of their nonparetic leg (**Figure 4B)**. The direction specificity of cortical-kinetic relationships is in line with the context-specific nature of cortically-mediated balance control,^33^ that may have less flexibility to adapt to changing environments (i.e., cortical engagement may reflect differing neural processes during paretic-loaded recovery, yielding less effective kinetic responses). These cortical-kinetic relationships provide an individualized framework for the clinical assessment and treatment of post-stroke balance impairment, showing individual differences in the nature and degree of impairment that can potentially be used to assess individuals, prescribe, and track treatment.

#### Conclusions

Our well-controlled reactive balance paradigm revealed that stroke-induced lesions may lead to slower and smaller cortical responses compared to age-matched controls, and illustrates a link between compromised engagement of cortical resources and post-stroke balance dysfunction. Specifically, our results suggest that individuals after stroke may be uniquely limited in their balance ability by slower cortical engagement, particularly under challenging balance conditions that rely on the paretic leg. These findings highlight the potential of temporal features of evoked cortical N1 responses to provide a biomarker of clinically-relevant balance and mobility behavior, offering a possible targeted avenue for rehabilitation efforts during stroke recovery.

## Data Availability

All data produced in the present study are available upon reasonable request to the authors.

## 5 Acknowledgements

We would like to acknowledge Michelle Goto, Pam Inglett, and Annie Goldman for their assistance with data collection for this project.

## 8. Supplementary material

None.

